# Preventive care in orthopaedic clinical services – testing the acceptability of an online health risk self-assessment tool using a multi-method design

**DOI:** 10.64898/2026.04.09.26350435

**Authors:** Simon R.E. Davidson, Stuart Browne, Luke Giles, Karen Gillham, Robin Haskins, Elizabeth Campbell, the Online Tool Working Group, Joseph Turner, Hayden Lowth, Lisa Corbett, Louise Robin, Kua Swan, Christopher M Williams, Emma Mudd

## Abstract

**Background:** Musculoskeletal conditions, such as back pain and osteoarthritis, are common and disabling disorders. Musculoskeletal conditions are closely related to chronic disease risk factors like smoking/vaping, poor nutrition, alcohol misuse and physical inactivity and impact a person’s risk of falling (SNAPF). Preventive care for SNAPF risks is often overlooked. Online delivery of preventive care may increase the provision of this care. We aimed to assess if an online tool for SNAPF risks would be used by and acceptable to patients waiting for an orthopaedic consultation.

**Methods:** We completed a multi-method study to test an online health risk self-assessment tool. A random sample of 300 people on the orthopaedic outpatient waiting list aged 18-64 years were sent the tool in batches of 20-50. The tool assessed SNAPF risks and provided feedback against national guidelines. After each batch, we completed feedback interviews with participants to assess acceptability and updated the tool. We summarised quantitative data using descriptive statistics and qualitative data using thematic analysis.

**Results:** Of the 300 participants sent the tool, 51.3% were female, 8.6% identified as Aboriginal and/or Torres Strait Islander, with a mean (SD) age of 52.0 years (11.2). There were 170 participants (59.2%) who completed the tool, 117 who did not complete it, and 13 participants who were excluded from analysis because they did not receive the SMS. We conducted 184 feedback interviews, including 125 ‘completers’ and 59 ‘non-completers’. The percentage of participants who felt that SMS was an appropriate way to receive the tool was 84.7% of ‘completers’ and 50% of ‘non-completers’. The two most common reasons for not completing the tool were due to perceived risk (13/59, 22.0%), and the SMS was received at an inconvenient time (11/59, 18.6%). Qualitative data from the feedback interviews captured three enablers: i) design, ii) high importance, and iii) engagement with health service, along with four barriers: i) design, ii) risk, iii) relevance, and iv) engagement with health service.

**Conclusion:** Our study found that an online health risk self-assessment tool appears to be an acceptable way to assess chronic disease and falls risk factors for people on an orthopaedic waitlist.

**Trial registration:** Not applicable.

## Background

Musculoskeletal (MSK) conditions, such as low back pain and osteoarthritis, are common in Australia and internationally.(1, 2) These conditions frequently co-occur with non-communicable diseases,(3, 4) placing high burdens on the health system, society and individuals.(3, 5) In 2020-21, MSK conditions were responsible for an estimated $14.7 billion of expenditure in the Australian health system and represented the highest spending of all disease groups. In 2018, 16% of the total burden of MSK conditions was attributed to modifiable chronic disease risk factors, such as overweight, obesity and tobacco use.(1)

High rates of modifiable chronic disease risks like smoking, poor nutrition, alcohol misuse, physical inactivity, and modifiable falls risk factors (SNAPF) are present in many people accessing the Australian health system. A study describing the rates of chronic disease risks in people awaiting orthopaedic consultation for a MSK condition found that 28% were current smokers, over 80% did not meet recommendations for fruit and vegetable intake, just under 20% overconsumed alcohol, more than 80% were physically inactive, and three-quarters had three or more modifiable risk factors. Only approximately half of these people reported having been advised about the benefits of addressing their modifiable risks.(6) While preventive care for chronic disease risks in clinical settings is recommended(7), it is often overlooked.(8, 9)

The lack of preventive care provided to people awaiting orthopaedic consultation represents an opportunity to explore the most effective and scalable way to deliver preventive care within this setting. Models of care such as the Ask, Advise and Help (AAH) model(10) provide an evidence-based way of structuring a clinical conversation about health behaviours that can be applied to modifiable chronic disease risks. Free government-funded population-level support services, such as New South Wales (NSW) Quitline (telephone support for smoking/vaping) and Get Healthy Service (GHS) (telephone and online support for healthy eating, healthy weight, physical activity and appropriate alcohol use) provide a widely accessible, effective(11, 12) referral options that can be delivered within the ‘Help’ component of the AAH model. Unfortunately, a common clinician-reported barrier to the delivery of preventive care in clinical appointments is limited time.(13) Digital health interventions, including online tools that assess risk factors and provide health advice and suggest support services, can potentially help address this gap in preventive care, complementing care provided in clinical consultations.(14)

We aimed to assess if an online health risk self-assessment tool for SNAPF risks would be used by patients waiting for an orthopaedic specialist consultation at a major public hospital and if it is an acceptable way to provide preventive care using the AAH model, from a patient perspective. Our secondary aims were to: i) describe the prevalence of SNAPF risk factors in this waitlist population, ii) explore whether people with MSK conditions value preventive care, and iii) explore whether people with MSK conditions were aware of and interested in referral to population-level support services for SNAPF risks.

## Methods

### Study design

We completed a multi-method study, using quantitative and qualitative data and a user-centred approach to test and improve an online health risk self-assessment tool. The Hunter New England Local Health District (HNELHD) Human Research Ethics Committee (2019/ETH01034) granted ethics approval. Clinical trial number: not applicable.

Our study had two phases.

- Phase 1- Initial scoping of acceptability involving informal face-to-face interviews with orthopaedic patients (March 2023)
- Phase 2- Batch testing and refinement of the online health risk self-assessment tool with a random sample of 300 patients awaiting orthopaedic consultation – eight batches of 20-50 patients (June 2023 - May 2024)

### Online health risk self-assessment tool development

We developed an online health risk self-assessment tool based on the AAH model of care for SNAPF risk factors (see Appendix One for the final version of the online tool). The tool was designed to include ‘Ask’ and simple ‘Advice’ that encouraged behaviour change and reinforced positive behaviour. ‘Advice’ was delivered across all test batches via a brief summary of the person’s SNAPF risks. Risk status was determined using Australian national guidelines for fruit and vegetable consumption,(15) physical activity,(16) alcohol intake,(17) and smoking/vaping.(18) Falls risk was assessed using a Local Health District (LHD) screening tool that considers age, falls in the last 12 months, number of prescribed medications, history of stroke or Parkinson’s disease, self-reported problems with balance and the need to use arms to get up from a chair. From batch four onwards, participants also had the option to view four health risk information sheets: 1) smoking, 2) physical activity, 3) nutrition and alcohol, and 4) falls. The information sheets were embedded into the tool for patients to obtain further information about SNAPF risks in relation to musculoskeletal conditions, pain, self-management, and included information on and links to support service websites (e.g., NSW Quitline and GHS). We built the online health risk self-assessment tool as a survey in the Research Electronic Data Capture program (REDCap).

### Setting

Our study was completed at a large tertiary referral hospital (899 beds) in a LHD in NSW, Australia. The orthopaedic outpatient department receives approximately 7,000 referrals annually. The LHD services 942,374 people and has a catchment area of 131,785km^2^, which includes metropolitan, inner and outer regional areas.(19) People on the orthopaedic outpatient department waiting list are most often referred for back or knee pain, and are waiting for an initial appointment with an orthopaedic specialist.(6)

### Participants and eligibility

#### Phase 1

Any patient who attended an appointment at the orthopaedic outpatient department on the four days the research team were visiting the site, and who agreed to an informal face-to-face interview.

#### Phase 2

A random sample of 300 patients on the orthopaedic outpatient department waiting list who had been referred with a hip, knee or spinal orthopaedic condition and were aged between 18 and 64 years.

### Testing procedures

#### Phase 1

Members of the research team (SD, EM and SB) approached orthopaedic patients in the waiting room of the outpatient department and asked if they would like to participate in an informal interview regarding an online tool to assess chronic disease risk factors. We asked patients whether they would open and complete an online tool sent to them via a link in a Short Message Service (SMS) from the hospital. We also showed them a draft design of the online tool and asked for feedback on the tool questions and risk factor summary page.

#### Phase 2

We undertook eight batch tests with 20-50 patients in each batch (see Figure 1). Participants were sent the online health risk self-assessment tool link via SMS and provided e-consent within the tool. Those participants who had not yet completed the tool after one week received a single follow-up reminder SMS. We performed an initial round of testing (batch 1) to confirm whether patients would open the SMS and follow a link to the tool. In batches 2-8, we used a split testing method (A/B testing) to test a range of tool design scenarios (e.g., SMS alpha tag branding, question format, length of tool, risk factor summary content, links to additional information sheets, and sending information sheets via SMS/email). Following each batch test, we reviewed survey analytics (e.g., completion rates) and undertook consumer feedback interviews with participants. Based on data analytics and feedback interviews, we made changes to the tool prior to the next batch test.

**Figure 1:**
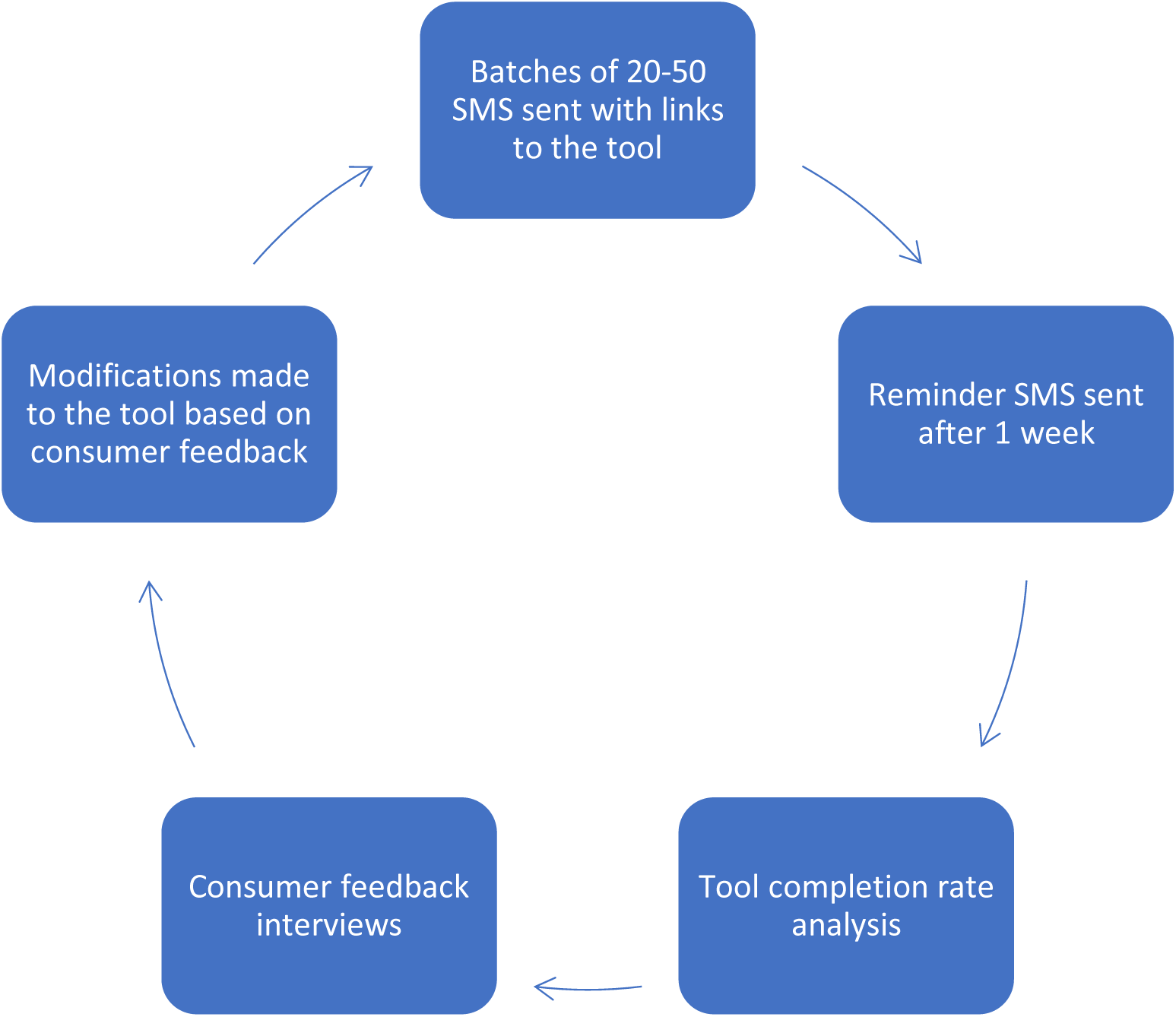
Cycle of batch testing the online health risk self-assessment tool

#### Consumer feedback interviews

We attempted to complete semi-structured feedback interviews with all participants who were sent the online health risk self-assessment tool and for whom we did not receive a delivery failure notification, regardless of whether they completed the tool or not. Telephone interviews commenced within 1-4 days following tool completion, or approximately two weeks after the tool invitation SMS was sent for non-completers. Interview questions focused on exploring tool acceptability (identifying the barriers and enablers to tool completion), participant perceptions of the value of preventive care, and awareness of and interest in referral to population-level support services for SNAPF risks. Interviews were completed by female and male Project Officers from a NSW LHD Health Promotion Service who were involved in the development and testing of the tool. Interviewers had between 1-14 years of experience in health promotion. An Aboriginal Project Officer was available for calls if requested by participants. Interviewers made five call attempts across different days before deeming a participant to be uncontactable. We used multiple-choice and open-ended questions (see Appendix Two for interview guide) during feedback interviews. The data was entered into a REDCap data collection tool.

### Data sources

For our primary aim, we used tool analytics (quantitative) and consumer feedback interviews (qualitative) to assess use and acceptability. For our secondary aims, we used tool responses (quantitative) to identify SNAPF prevalence, and data from consumer feedback interviews to explore the perceived value of preventive care for people with MSK conditions and the awareness of and interest in referrals to population-level support services (quantitative).

### Statistical Analysis

We summarised quantitative data using descriptive statistics, either mean and standard deviation (SD) or median and interquartile range (IQR), depending on distribution. We summarised categorical variables using n (%). We categorised the descriptive statistics into two groups: i) <u>Completers</u>- participants who fully completed the online health risk self-assessment tool and ii) <u>Non-completers</u>- participants who did not open the tool or opened the tool but did not consent or did not fully complete the tool.

The interviewers who undertook most consumer feedback interviews (SB & LG) analysed the qualitative data. We took a pragmatic approach to the thematic analysis(20) of feedback to identify common themes for each question following each batch of testing. The interviewer used a codebook of previously identified themes from previous test batches, but also added new relevant themes to the codebook as they were identified through the different batch tests.

## Results

### Phase 1

We undertook 10 face-to-face interviews with in-scope patients waiting to see a consultant in the orthopaedic outpatient department. Almost all participants reported that they would open and complete an online tool that was sent via SMS from the hospital. They also reported that the SMS invitation wording and online health risk self-assessment tool questions and summary concept (based on relevant national guidelines) were acceptable. Overall, phase 1 scoping interviews indicated support for the online health risk self-assessment tool concept, therefore we progressed to Phase 2.

### Phase 2- Sample characteristics

Of the 300 participants sent the tool, 51.3% were female, 8.6% identified as Aboriginal and/or Torres Strait Islander, and participants had a mean (SD) age of 52.0 years (11.2). The two most common conditions for which patients were referred to the outpatient service were knee pain (63.3%) and hip pain (19.3%). Most participants were triaged to be seen within 365 days (78.3%). We received an SMS delivery failure notification from 13 participants (4.3%), who were excluded from further analysis.

We attempted to contact all participants who received the online health risk self-assessment tool (n=287) for consumer feedback interviews. Of the 287 participants, we were able to contact 225 (78.4%), and 184 (64.1%) agreed to provide feedback. Consumer feedback participants had a mean (SD) age of 52.8 years (10.5), 51.1% were female, and 9.7% identified as Aboriginal and/or Torres Strait Islander.

### Primary Aim

#### Quantitative data

There were 287 participants included in the analysis who received the invitation SMS. Of those 170 participants (59.2%) fully completed the tool. The remaining 117 participants were categorised as non-completers and included eight (2.8%) partial completions, 14 (4.9%) who opened the tool but did not complete any items, and 95 (33.1%) who did not open the tool. Of those who participated in interviews, 125 participants had completed the tool (representing 73.5% of ‘completers’), and 59 had not completed the tool (representing 50.4% of ‘non-completers’).

The consumer feedback interview data showed most ‘completers’ felt that SMS was an appropriate way to receive the online health risk self-assessment tool (84.7%), while this was lower for the ‘non-completers’ (50.0%). The three most common reasons for not completing the tool were that participants deemed it too risky (13/59, 22.0%), the SMS was received at a non-convenient time (11/59, 18.6%), and their orthopaedic condition had resolved and the participant no longer required care (9/59, 15.3%). Most participants in both the ‘completers’ (92/124, 74.2%) and ‘non-completers’ (29/40, 72.5%) groups felt it was positive that the health service was asking about chronic disease risk factors. Table 1 shows additional variables relating to the use and acceptability of the tool components, including the potential addition of advice and referral (asked in early batches).

**Table 1:**
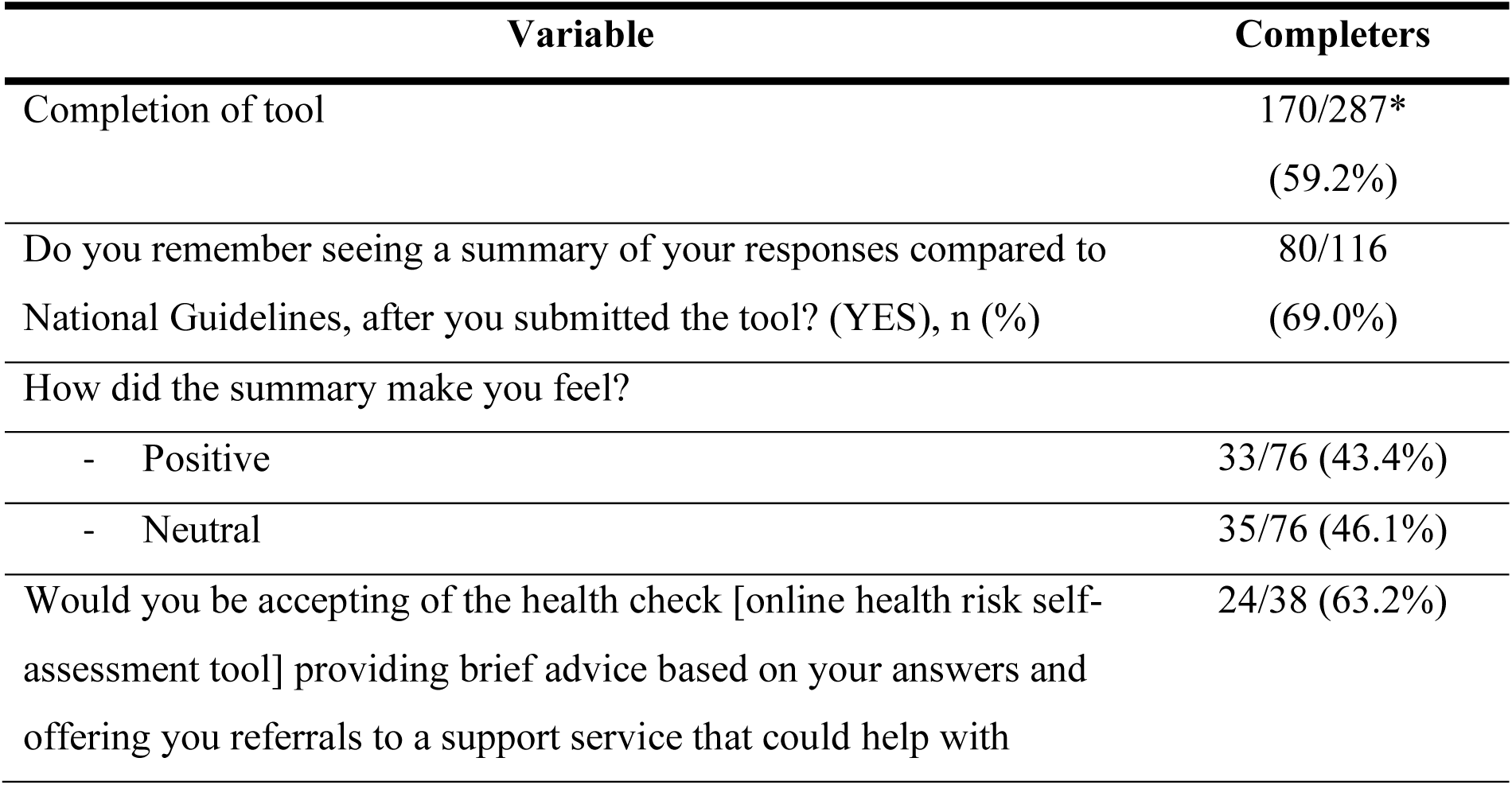
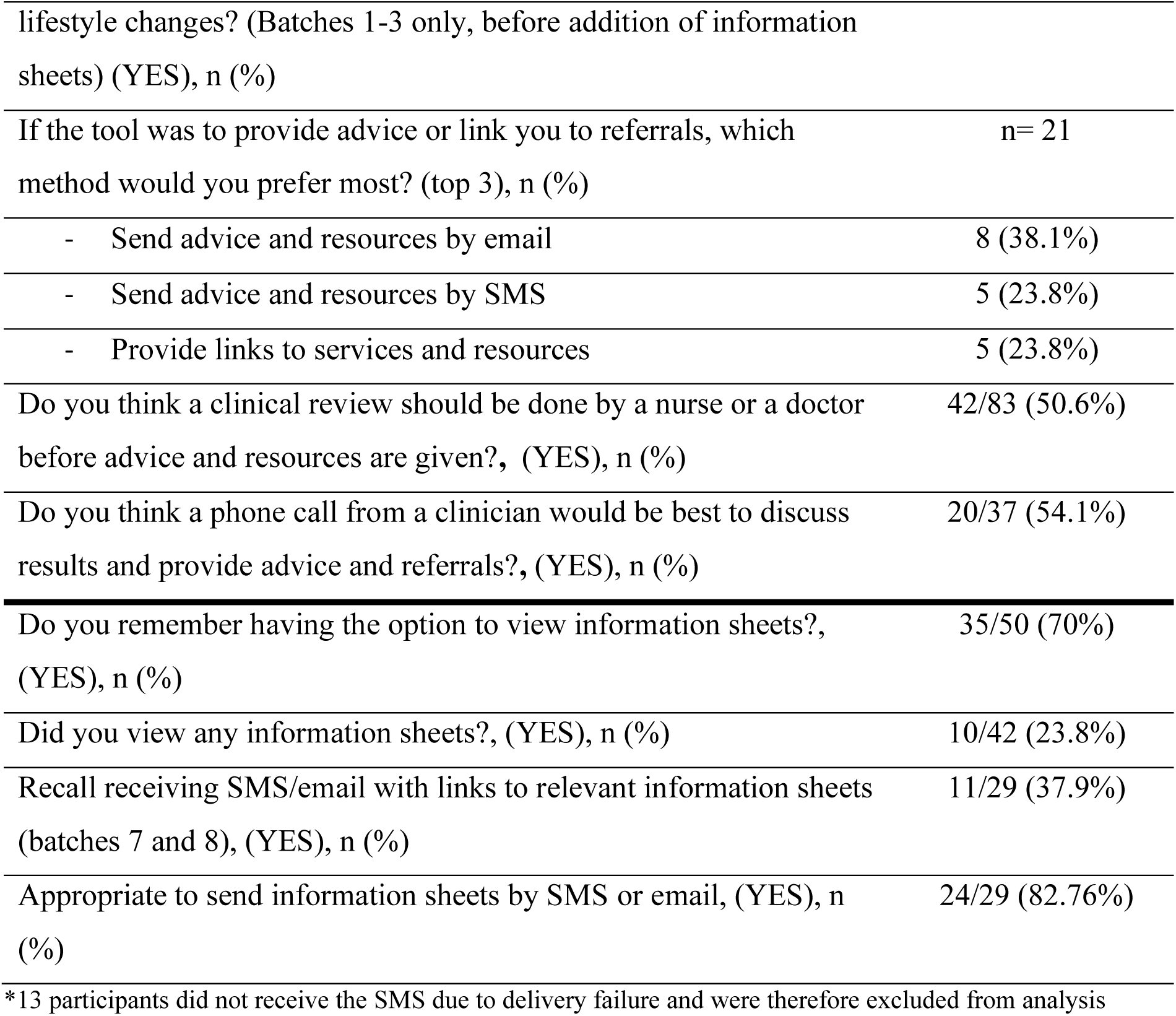
Use and acceptability of online health risk self-assessment tool components.

#### Qualitative data

Qualitative data from the consumer feedback interviews captured a mix of positive (enablers) and negative (barriers) themes. See Figure 2 for descriptions of the identified enablers and barriers. Three key enablers were identified: i) design, ii) high importance, and iii) engagement with health service. Elements of the online health risk self-assessment tool, like the user-friendly design, as well as participants’ perceived importance of chronic disease risks, and contact from the health service during waiting times informed these positive themes.

**Figure 2:**
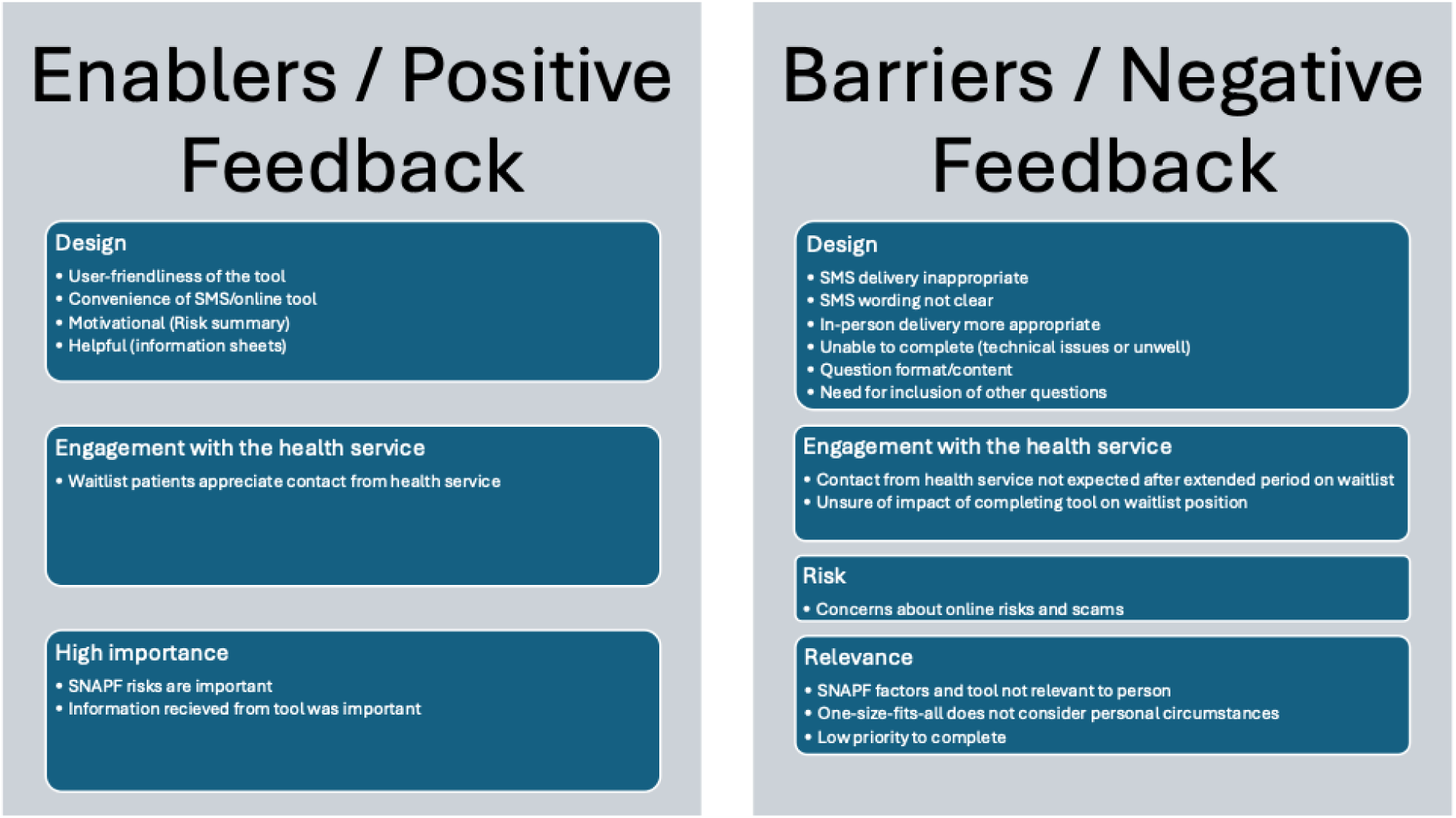
Qualitative themes from consumer feedback interviews. Note: SMS= Short Message Service, SNAPF= Smoking, Nutrition, Alcohol, Physical Activity, Falls

In contrast, four barriers were identified: i) design, ii) risk, iii) relevance, and iv) engagement with the health service. Concerns about online security risk, the one-size-fits-all approach, and participants’ preference for preventive care to be delivered face-to-face were some of the elements that informed these themes. Two themes emerged that were both an enabler and a barrier. The first theme was ‘design’. Elements of the design like the user-friendliness and the convenience were reported as positive elements; however, some participants felt the design was inappropriate for the care that was being delivered and preferred alternative methods to discuss chronic disease risks and receive information (e.g., face-to-face, email). The second theme was the ‘engagement with health service’; some participants appreciated contact from the health service while on the outpatient waiting list, while others were not expecting the contact and did not know what effect completing the tool (and their answers) would have on their progress on the waiting list. More detail regarding the qualitative data from consumer feedback interviews are provided in Appendix Three.

### Secondary Aims

#### Phase 2- SNAPF risk prevalence

Participants who completed the tool had a high prevalence of SNAPF factors (Table 2). Only five participants (2.9%) had no SNAP risks, while 71.8% had two or more risks. Participants aged from 50-64 years were also asked about their falls risk, with over one-third (38.6%) of participants identified as being at high risk of falls.

**Table 2:** Proportion of ‘completers’ with SNAPF risk factors.

| **Nutrition\*** | **n (%)** |
| --- | --- |
| - Low fruit intake (<2 serves/day) | 112/170 (65.9%) |
| - Low vegetable intake (<5 serves/day) | 154/170 (90.6%) |
| **Physical inactivity\*\*** | 64/170 (37.7%) |
| **Excess alcohol intake** | 72/170 (42.4%) |
| **Smoking status** |  |
| - Never smoked | 58/170 (34.1%) |
| - Ex-smoker | 67/170 (39.4%) |
| - Current smoker | 45/170 (26.5%) |
| - Smoking (in the last 4 weeks)*** | 46/112 (41.1%) |
| **E-cigarette use (Vaping)** |  |
| - Never used e-cigarettes | 95/110 (86.4%) |
| - Ex-e-cigarette user | 4/110 (3.6%) |
| - Occasional e-cigarette user | 5/110 (4.5%) |
| - Daily e-cigarette user | 6/110 (5.5%) |
| **Total number of risk factors (falls excluded)** |  |
| - 0 risks | 5/170 (2.9%) |
| - 1 risk | 43/170 (25.3%) |
| - 2 risks | 76/170 (44.7%) |
| - 3 risks | 36/170 (21.2%) |
| - 4 risks | 10/170 (5.9%) |
| **Falls** |  |
| - 50 years and over (at time of completion) | 132/170 (77.6%) |
| - Fall in the last 12 months | 42/132 (31.8%) |
| - 4 or more prescribed medications a day | 62/132 (47.0%) |
| - Diagnosed with cerebrovascular accident (stroke) or Parkinson's disease | 4/132 (3.0%) |
| - Balance problems (unsteadiness, dizziness) | 65/132 (49.2%) |
| - Need to use arms to get up from a chair | 92/132 (69.7%) |
| **Falls risk status** (age 50-65 only, n=132) |  |
| - No risk (0 risk factors) | 25/132 (18.9%) |
| - Low risk (1-2 risk factors) | 56/132 (42.4%) |
| - High risk (>2 risk factors) | 51/132 (38.6%) |

\* overall nutrition risk was determined by combining questions regarding fruit and vegetable intake (i.e. participants not meeting fruit and/or vegetable intake recommendations were classified as at risk for nutrition).
\*\*responses >30 hours per week were truncated to a value of 30 hours per week, to reflect more probable physical activity levels. This impacted on 5 responses, but did not change the percentage of participants meeting physical activity guidelines.
\*\*\*question asked of current smokers and ex-smokers only.

#### Perceived value of preventive care for people with MSK conditions

When asked about the perceived importance of individual SNAPF risks, both ‘completers’ and ‘non-completers’ rated the importance as at least 7/10 (0 being not important at all, 10 being most important) for all SNAPF risks. Most people (92.0%) felt that it was important that health staff knew about SNAPF risks. Over a quarter of completers (27.8%) and 39.5% of non-completers had not discussed their SNAPF risks with a health professional in the last year. See Table 3 for further details.

**Table 3:** Perceived importance of chronic disease risks.

| Variable | Completers | Non-completers |
| --- | --- | --- |
| <u>Perceived importance of chronic disease risks and health service clinicians being aware (0-10), mean (SD) [n]</u> |  |  |
| How important is it to you to not smoke, reduce smoking or quit smoking? | 7.8 (3.6)<br>[n=107] | 8.5 (3.0)<br>[n=34] |
| How important is it to you to maintain a healthy and balanced diet? | 8.4 (1.7)<br>[n=112] | 8.2 (1.8)<br>[n=36] |
| How important is it to you to maintain a healthy alcohol consumption? | 7.3 (3.2)<br>[n=104] | 7.7 (2.8)<br>[n=35] |
| How important is it to you to maintain an active lifestyle and exercise regularly? | 8.4 (1.6)<br>[n=108] | 7.9 (2.3)<br>[n=36] |
| How important is it to you to maintain strength and balance – to reduce the risk of having a fall and injuring yourself? | 9.1 (1.2)<br>[n=105] | 8.8 (1.5)<br>[n=35] |
| Do you think it is important that health staff know about these chronic disease risk factors? Yes, n (%) | 103<br>(91.2%)<br>[n=113] | 35<br>(94.6)<br>[n=37] |

#### Awareness of and interest in referral to population-level support services

Slightly less than half (48.7%) of participants were aware of the GHS, while 78.4% of participants were aware of Quitline. Only one person (1/148, 0.7%) had used the GHS, 9.6% (7/73 smokers/ex-smokers) had ever used the Quitline phone service, and 3.4% (5/148) of participants had used a different telephone support service. When asked in Batch 1-3 if participants would be accepting of the online tool providing brief advice and referrals to a support service that could help with lifestyle changes, 60.0% (24/40) reported they would be. However, when asked in Batch 7 and 8 if participants would be interested in referral to specific support services (GHS and/or Quitline), only 30.8% (12/39) indicated they were.

## Discussion

We describe the use and acceptability of an online health risk self-assessment tool to provide preventive care for people on a public hospital orthopaedic outpatient waiting list. The tool appears to be acceptable to many in this patient group, with most able to receive an SMS with a link to the tool and over half (59.2%) of the participants fully completing the tool. Participants felt that it was important for health staff to know about SNAPF risks and found the risk assessment summary to be positive. However, there were mixed views on whether SMS delivery of the tool was appropriate (84.7% of ‘completers’ felt SMS was appropriate, compared to 50.0% of ‘non-completers’), negative views were largely due to the concerns relating to the risk of following hyperlinks. SNAPF risks were highly prevalent in the studied orthopaedic population, with over 70% of participants having two or more modifiable chronic disease risks. Both completers and non-completers of the tool rated the importance of addressing SNAPF risks as high (7+/10) and indicated that clinicians should be aware of their patients’ risks. Participant awareness of population-level support services that could be used to address SNAPF risks and desire for referral to these services varied.

Preventive care is an integral part of the Australian health system. The assessment and management of SNAPF risks is an important component of preventive care. Digital health interventions provide an opportunity to deliver this care at a convenient time for the consumer and reduce the burden on the clinician during appointments.(21) Our tool addresses a current gap within the digital health literature of tools that deliver preventive care by assessing multiple modifiable risks, including chronic disease and falls risks. A review of digital behavioural interventions for SNAP risks and obesity in young adults found that only three of 45 addressed multiple risks.(22) As well as this, many current digital tools do not provide all components of the AAH model, and often only concentrate on a single component (predominantly ‘Help’), aiming to bring about behaviour change, which may reduce their effectiveness.(21, 23) Our study shows that within an online preventive care tool, the ‘Ask’ component is received by approximately half of the participants. Most participants looked at the risk factor summary page (‘Advise’) and found it useful; however, when more action (e.g. additional clicks) was required from participants to access further information (via information sheets), these numbers reduced considerably. Finally, while participants were provided with referral links for population-level telephone support services, we do not know how many accessed these links.

The high prevalence of SNAPF risks in our study aligns with a previous study in orthopaedic outpatient services.(6) However, no studies have described the prevalence of SNAPF risks in other outpatient specialties. While participants felt SNAPF risks were important, many (27.8%-39.5%) had not discussed them with a health professional in the last year, perhaps indicating the patients or their health professional did not prioritise this care. Finally, the variable knowledge of population-level telephone support services for SNAPF risks identified in our study indicates that these services may need to be promoted more to specific patient groups to increase referrals to, and engagement with, the services. While awareness of Quitline (for smoking cessation) was high, the majority of patients were not aware of the GHS supporting healthy eating, physical activity, and healthy weight. These findings support the likely value in further efforts to increase preventive care for SNAPF in the orthopaedic outpatient setting and of further exploring the role of population-level support services as a ‘Help’ component.

### Strengths and Limitations

Our study has several strengths. We used a user-centred approach that gave participants the opportunity to provide feedback on their use of the tool. We tested the tool with a large number of people (n=287) and undertook consumer feedback interviews with 184 participants, giving us confidence in our findings. We also adapted the tool after each round of batch testing and feedback interviews, which allowed us to be receptive to findings and continually test the tool to develop an optimal version. A final strength of our study is the inclusion of falls risk screening in those aged 50 years or over, which is important to consider with the global ageing population and the health care costs associated with falls.(24)

Our study has a number of limitations. Participants on the waiting list often wait for extended periods and were not expecting to receive the SMS with the link to the online health risk self-assessment tool. Therefore, participants may have associated their completion of the tool with a faster orthopaedic appointment, and thus, our completion rates, clinical contexts or acceptability findings may not generalise to other contexts or clinical settings (e.g., orthopaedic settings where an appointment is pending, or community services). The tool does not consider any modifications to SNAPF risk assessment, advice or help based on specific health conditions (e.g., pregnancy). Nor was there an in-depth exploration of the delivery of the ‘Advise’ or ‘Help’ component of the AAH model via the digital tool. However, as we were undertaking initial acceptability testing, these adaptations can be made in future versions of the tool.

### Clinical implications

The online health risk self-assessment tool appears to be an acceptable digital solution to deliver brief preventive care. Clinical services can use the tool to deliver preventive care to patients on appointment waiting lists or with upcoming appointments. This will encourage these patients to be aware of their SNAPF risk profile, and provide the clinician with the opportunity to use the tool results and the clinical appointment to encourage and support positive behaviour change. While a digital tool may never reach all patients, if it is integrated into care pathways, it may be used by a large proportion of the target population and could save clinicians time during appointments by undertaking AAH components of preventive care. Identifying and supporting patients to address relevant SNAPF risks early may also reduce the risk of developing chronic diseases and falls, and reduce the health care required for a person’s chronic disease or fall-related injuries.

Many health services are currently transitioning to electronic health records. For example, NSW Health in Australia is planning to roll out a Single Digital Patient Record across the state between 2026 and 2028 to support its over eight million residents. Digital tools, like the online health risk self-assessment tool that we tested, may be integrated into electronic health records to provide preventive care to large numbers of patients at a low cost to the health service, potentially producing significant health improvements within the general population.

### Future Research

Future research should assess the acceptability of integrating advice and self-referral options into future iterations of the online health risk self-assessment tool. Research should also investigate the effectiveness of digital tools that include all components of the AAH model, and the timing of their delivery, to make positive impacts on modifiable chronic disease risks including SNAPF, and the effects these changes have on patient health and health service use. Finally, future research should also explore the capacity of digital tools to prompt self-referral to population-level support services and how digital preventive care tools can be integrated into clinical services to ensure they improve the care pathway for both patients and clinicians.

### Conclusion

Our study found that an online health risk self-assessment tool was an acceptable way to assess chronic disease and falls risk factors in patients on an orthopaedic waitlist. We found that SNAPF risk factors were high in this client group, and many clients were accepting of the tool containing brief advice and referral options to support services such as Quitline and GHS. Future research should explore whether preventive care delivered via digital tools can facilitate self-referral to population-level support services, and if it results in behaviour change and improvements in health outcomes.

## Supporting information

Appendix One

Appendix Two

Appendix Three

## Data Availability

Data used for analysis will be made available on reasonable request. Proposals for data use may be submitted to the principal/corresponding author.

## List of abbreviations

MSK: Musculoskeletal
SNAPF: smoking, poor nutrition, alcohol misuse, physical inactivity, and modifiable falls risk factors
AAH: Ask, Advise and Help
NSW: New South Wales
GHS: Get Healthy Service
HNELHD: Hunter New England Local Health District
LHD: Local Health District
REDCap: Research Electronic Data Capture program
SMS: Short Message Service
SD: Standard deviation IQR-Interquartile range

## Declarations

### Ethics approval and consent to participate

The study was approved by the Hunter New England Local Health District (HNELHD) Human Research Ethics Committee (Approval No. 2022/ETH02425). The study was completed in accordance with the Declaration of Helsinki. All participants provided electronic informed consent before completing the online tool and verbal informed consent prior to participating in a consumer feedback interview.

### Consent for publication

Not applicable.

### Authors Contributions

SD, SB, and EC contributed to the design of the study and development of the online health risk self-assessment tool. KG and RH provided operational support and guidance. SD coordinated the data collection and analysis. SB and LG completed the data collection and analysis. SD and SB prepared the initial draft of the manuscript. All authors provided critical appraisal and approved the final version. All listed authors meet authorship criteria, and no others that meet the criteria have been omitted.

## Acknowledgements

Online Tool Working Group members:

Joseph Turner^4^, Hayden Lowth^4^, Lisa Corbett^4^, Louise Robin^1^, Kua Swan^1^, Christopher M Williams ^2^, and Emma Mudd^2^.

