## Appendix One for "Preventive care in orthopaedic clinical services – testing the acceptability of an online health risk self-assessment tool using a multi-method design"

### HNELHD Lifestyle Health Check

Your lifestyle may affect your overall health, joint pain, recovery and outcomes of surgery. Smoking, low fruit and vegetable consumption, physical inactivity or drinking above the recommended amount of alcohol may also increase your risk of developing chronic diseases.

Identifying and making small changes to your lifestyle (where possible) may improve your health and help you manage pain better. Please take time to answer the following health-related lifestyle questions. Completion is non-compulsory, will take approximately 2-5 minutes and will not affect your timeline to an appointment. On completion a short summary of your answers compared to Australian national guidelines will be provided along with further relevant information.  We encourage you to read the information provided and speak with your doctor or health professional at your next visit to review your answers and to determine if there are appropriate steps you could take to improve your health.

---

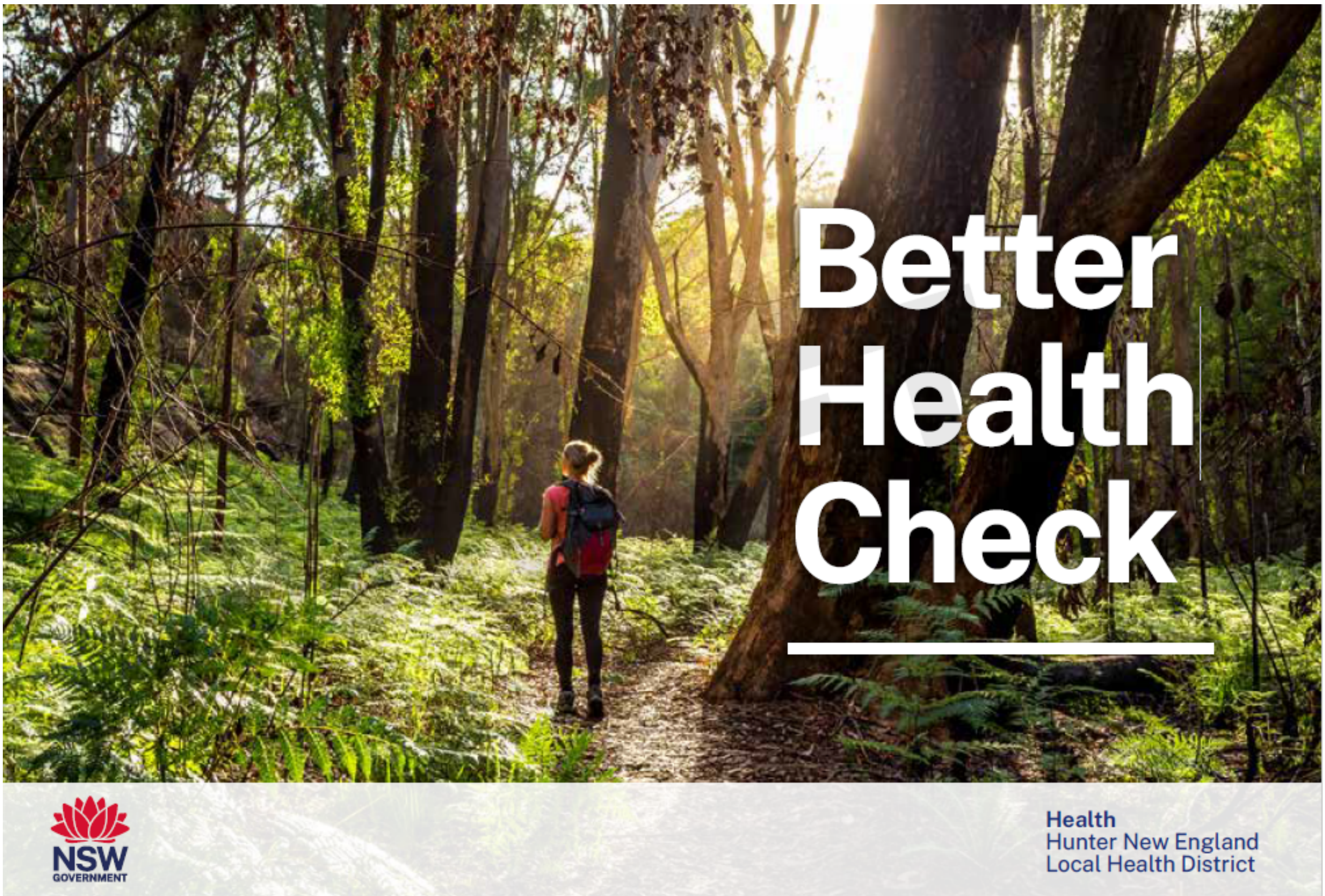


---

Please answer the health-related lifestyle questions below.

Are you happy to continue?

◯ Yes
◯ No

#### Your details

| | |
|---|---|
| First name | ________________ |
| Surname | ________________ |
| Do you identify as Aboriginal or Torres Strait Islander? | ◯ No<br>◯ Yes<br>◯ Prefer not to say |
| If yes, do you identify as: | ◯ Aboriginal<br>◯ Torres Strait Islander<br>◯ Both |
| 6 Are you currently pregnant or planning a pregnancy? | ◯ No<br>◯ Yes, pregnant<br>◯ Yes, planning a pregnancy |
| Email address<br><br>Note: your email address will only be used for sending you links to relevant health information sheets based on your responses to the health-related lifestyle questions. | ________________ |

#### Fruit and vegetable intake

| | | |
|---|---|---|
| 1 | How many serves of fruit do you eat each day? | ◯ 0<br>◯ 1<br>◯ 2 or more<br>(A serve is 1 medium piece or 1 cup of chopped/canned fruit) |
| 2 | How many serves of vegetables do you eat each day? | ◯ 0<br>◯ 1<br>◯ 2<br>◯ 3<br>◯ 4<br>◯ 5 or more<br>(A serve is approximately 1 cup of salad or 1/2 a cup of cooked vegetables) |

#### Physical activity levels

4 How many hours per week do you spend doing moderate physical activity? (e.g. such as a brisk walk, golf, mowing the lawn or swimming)

For example, for 1 hour and 30 minutes of moderate physical activity per week, enter as 1.5, etc.

________________________________
(Moderate physical activity means you can still talk comfortably, but are unable to sing OR approximately 3-7/10 effort.)

3 How many hours per week do you spend doing vigorous physical activity? (e.g. jogging, aerobics, fast cycling, soccer or netball)

For example, for 1 hour and 30 minutes of vigorous physical activity per week, enter as 1.5, etc.

________________________________
(Vigorous activity means you can't talk comfortably, and huff and puff (7-10/10 effort))

Total number of hours activity

________________________________

#### Alcohol intake

| | | |
|---|---|---|
| 5 | How often do you have a drink containing alcohol? | ◯ Never<br>◯ Monthly or Less<br>◯ 2 to 4 times a month<br>◯ 2 to 3 times a week<br>◯ 4 or more times a week |
| 5.1 | How many drinks containing alcohol (standard drinks) do you have on a typical day when you are drinking? | ◯ 1 or 2<br>◯ 3 or 4<br>◯ 5 or 6<br>◯ 7 to 9<br>◯ 10 or more |
| 5.2 | How often do you have five or more standard drinks on one occasion OR more than 10 standard drinks in a week? | ◯ Never<br>◯ Less than monthly<br>◯ Monthly<br>◯ Weekly<br>◯ Daily or almost daily |
| | Total score | ______________________________ |

#### Smoking Habits

| | | |
|---|---|---|
| 7 | Which best describes your tobacco use/smoking (including all forms e.g. cigarettes, cigars, pipes, Shisha)? | ◯ Current smoker<br>◯ Ex-smoker<br>◯ Never been a smoker |
| | How often do you smoke tobacco? | ◯ Daily smoker<br>◯ Occasional smoker (less than daily) |
| 7.1 | When did you stop smoking? | __________________________________ |
| 7.2 | Have you smoked any tobacco in the past 4 weeks? | ◯ Yes<br>◯ No |
| 7.3 | What form/s of tobacco did you smoke? | ☐ Cigarette<br>☐ Roll your own<br>☐ Cigar<br>☐ Water pipe/Shisha/Hookah<br>☐ Tobacco mixed with cannabis<br>☐ Other (Free text answer) |
| 7.4 | Other form of tobacco smoked: | __________________________________ |
| | Which best describes your e-cigarette/vape device use? | ◯ Never used e-cigarettes or vape device<br>◯ Daily e-cigarette/vape use<br>◯ Occasional e-cigarette/vape use<br>◯ Quit e-cigarette/vape use |
| | Have you used a e-cigarette/vape device in the last 4 weeks? | ◯ Yes<br>◯ No |

#### Falls

| | | |
|---|---|---|
| 8 | Are you 50 years old or over? | ◯ Yes<br>◯ No |
| 8.1 | Have you had a fall in the last 12 months? | ◯ Yes<br>◯ No<br>◯ Unknown |
| 8.2 | Do you take 4 or more prescribed medications a day? | ◯ Yes<br>◯ No<br>◯ Unknown |
| 8.3 | Have you ever had a stroke or do you have Parkinson's disease? | ◯ Yes<br>◯ No<br>◯ Unknown |
| 8.4 | Do you have problems with your balance? This could be feeling unsteady on your feet or dizzy and lightheaded at times. | ◯ Yes<br>◯ No<br>◯ Unknown |
| 8.5 | Do you need to use your arms to get up from a chair? | ◯ Yes<br>◯ No<br>◯ Unknown |
| | Please add any additional health or lifestyle information that you feel may be relevant. | ______________________________ |

#### Summary

Thank you for taking the time to complete the lifestyle health check. Your results based on national guidelines are outlined below.

When you have finished reviewing your summary and information links, please click the 'SUBMIT' button at the bottom of this page.

Nutrition risk (1=met, 0=not met) ______________________

PA risk (1=met, 0 not met) ______________________

Alcohol risk (1=met, 0=not met) ______________________

Smoking risk (1=met (non-smoker), 0=not met (smoker)) ______________________

Vaping risk (1=met, 0=not met) ______________________

PA info sheet calc text for SMS ______________________

Nutrition/alcohol info sheet calc text for SMS ______________________

Smoking/vaping info sheet calc text for SMS ______________________

Falls info sheet calc text for SMS ______________________

Nutrition You are not quite meeting the recommended intake of 2 serves of fruit and 5 serves of vegetables each day. If you have received specific advice on nutrition from a health professional, please continue to follow that advice.  For support to eat healthy, call the Get Healthy Service on 1300 806 258 or go to www.gethealthynsw.com.au.

Would you like more information on healthy eating and drinking, or available support services?
- ◯ Yes
- ◯ No

To view more information about nutrition and available support services please click the link below:

Healthy Eating and Drinking Habits

Nutrition Well done. You are currently meeting the national guidelines for fruit and vegetable intake. Keep it up!

Would you like more information on healthy eating and drinking, or available support services?
- ◯ Yes
- ◯ No

To view more information about nutrition and available support services please click the link below:

Healthy Eating and Drinking Habits

Physical Activity You are not quite meeting the recommended 30 minutes of moderate-intensity physical activity on most (5+ days), days of the week. If you have received specific advice on activity from a health professional, please continue to follow that advice. For support to get physically active, call the Get Healthy Service on 1300 806 258 or go to www.gethealthynsw.com.au.

Would you like more information on physical activity and available support services?
◯ Yes
◯ No

To view more information about physical activity and available support services please click the link below:

Exercise and your health

Physical Activity Well done for being active. You are currently meeting the national guidelines of doing at least 30 minutes of moderate-intensity physical activity on most (5+ days), days of the week. Keep it up!

Would you like more information on physical activity and available support services?
◯ Yes
◯ No

To view more information about physical activity and available support services please click the link below:

Exercise and your health

Alcohol If you are pregnant (or planning a pregnancy), you should not drink alcohol. The alcohol you drink at any stage of pregnancy passes directly to your baby and can damage their developing brain, body and organs. It is never too late to stop drinking alcohol during pregnancy. It may be difficult for you to stop drinking alcohol - if so, it is really important to seek help from a doctor, midwife, obstetrician or Aboriginal Health Worker. You can also call the Get Healthy Service on 1300 806 258  or go to www.gethealthynsw.com.au.

Alcohol Keep it up! If you are pregnant (or planning a pregnancy), you should not drink alcohol. The alcohol you drink at any stage of pregnancy passes directly to your baby and can damage their developing brain, body and organs.

Would you like more information on alcohol, nutrition and available support services?
◯ Yes
◯ No

To view more information about drinking alcohol and available support services, please click the link below:

Healthy Eating and Drinking Habits

Alcohol Great work. You are currently meeting the national guidelines to reduce health risks from drinking alcohol. Keep it up!

Would you like more information on healthy eating and drinking, or available support services?
◯ Yes
◯ No

To view more information about drinking alcohol and available support services, please click the link below:

Healthy Eating and Drinking Habits

Alcohol You may be drinking more than the recommended number of standard drinks per week (more than 4 on any one day or more than 10 in a week). For support to reduce alcohol, call the Get Healthy Service on 1300 806 258 or go to www.gethealthynsw.com.au.

Would you like more information on alcohol, nutrition and available support services?
◯ Yes
◯ No

To view more information about drinking alcohol and available support services, please click the link below:

Healthy Eating and Drinking Habits

Smoking It's recommended to quit smoking for your overall health. For support to quit smoking, speak with your health care provider, call the Quitline on 137848 or go to www.icanquit.com.au

Smoking Stopping smoking has many health benefits for you and your baby. For support to quit smoking, speak with your health care provider, call the Quitline on 137848 or go to www.icanquit.com.au

Would you like more information on smoking, vaping and your health, or available support services to quit?
◯ Yes
◯ No

To view more information about smoking and available support services, please click the link below:

Smoking - Joint Disease and Pain

Smoking Keep it up! Not smoking is one of the best things you can do for your health. For support to stay smoke free, speak with your health care provider, call the Quitline on 137848 or go to www.icanquit.com.au

Smoking Well done! Being smoke free has many health benefits for you and your baby. For support to stay smoke free, speak with your health care provider, call the Quitline on 137848 or go to www.icanquit.com.au

Smoking Well done, not smoking is one of the best things you can do for your health.

Would you like more information on smoking, vaping and your health, or available support services?
◯ Yes
◯ No

To view more information about smoking and available support services, please click the link below:

Smoking - Joint Disease and Pain

Vaping Every vape is a hit to your health. For support to quit vaping, speak with your health care provider, call the Quitline on 137848 or go to www.icanquit.com.au

Vaping Vaping is not safe for you or your baby. For support to quit vaping, speak with your health care provider, call the Quitline on 137848 or go to www.icanquit.com.au

Would you like more information on smoking, vaping and your health, or available support services to quit?
◯ Yes
◯ No

To view more information about smoking and vaping, and available support services, please click the link below:

Smoking & Vaping - Joint Disease and Pain

Vaping Well done, not vaping has many health benefits for you and your baby. For support to stay vape free, speak with your health care provider, call the Quitline on 137848 or go to www.icanquit.com.au

Vaping Well done, not vaping has many health benefits. For support to stay vape free, speak with your health care provider, call the Quitline on 137848 or go to www.icanquit.com.au

Vaping Well done, not vaping has many health benefits.

Vaping Well done, not vaping has many health benefits for you and your baby.

Would you like more information on smoking, vaping and your health, or available support services to quit?

- ◯ Yes
- ◯ No

To view more information about smoking and vaping, and available support services, please click the link below:

Smoking & Vaping - Joint Disease and Pain

Falls Having a fall can lead to a less independent lifestyle or serious injury. Your answers suggest that you may be at risk of a fall. It is recommended you seek further assessment with a health professional.

Falls Prevention Would you like more information about falls prevention and available support services?

- ◯ Yes
- ◯ No

Falls Prevention There are many factors that can contribute to your risk of having a fall and injuring yourself. To view more information about falls prevention and available support services, please click the link below:

Falls Prevention

Mail or Email Option If you would prefer to have information sheets mailed or emailed to you, please select which sheets you would like sent and provide either your postal or email address below.

- ☐ Healthy Eating and Drinking Habits
- ☐ Exercise and your health
- ☐ Smoking - Joint Disease and Pain (includes vaping)
- ☐ Falls Prevention

Please provide your postal or email address:

__________________________________

The results will be added to your medical record so that relevant HNELHD staff can review them as needed. Are you happy for that to occur?

- ◯ Yes
- ◯ No
