## Appendix Two for "Preventive care in orthopaedic clinical services – testing the acceptability of an online health risk self-assessment tool using a multi-method design"

### Appendix Two- Consumer feedback interview guide

1. Completer- Why did you complete the whole health check? (e.g. because the SMS said it was from HNELHD)
2. Non-completer- What were the reasons stopping you from (+ - fully) completing the health check? What was the main reason among those you have mentioned?
3. What are your thoughts on the health service asking you about these health and lifestyle factors?
4. If an explanation about SMS health check required, follow-up question about how the patient feels about being asked about those health lifestyle factors now?
5. On a scale of 0-10 (0 not important at all and 10 very important), how important are those lifestyle factors to you? Why that number? Why are they important?
   - If finished the health check – the health check had a summary at the end – did you have any thoughts on that section, either positive or negative – did you look at it, was it useful, could you see it on your device etc.
6. Have you discussed any of these lifestyle factors with your General Practitioner or other clinicians while you have been on the waiting list?
7. Are you aware of, or have you previously used phone support services such as Quitline and Get Healthy Service? (Quitline is a telephone support service to help quit smoking; Get Healthy Service is a telephone support service to help people make healthy lifestyle changes)
8. Do you have any questions or suggestions for how the health check could be improved?
   - Is SMS/online appropriate?
   - Could you read/see it ok (did it display properly on your phone or screen→ confirm type of phone) Explain answer.
   - Could you see the photo at the start of the questions? Do you have any thoughts on whether or not the photo was suitable for this type of health check?
   - Was further explanation about the health check required? Explain answer
   - Was the wording of the questions appropriate? Explain answer
   - Were there any questions that didn't make sense, or you were wondering why they were there?
   - If the health check was to provide brief advice based on your answers and offer you a referral to a support service that could help with lifestyle, would you be accepting of this?
     These services may include, for example free telephone support from NSW Quitline (for smoking), and the NSW Get Healthy Service; local services, such as group exercise programs, Aboriginal Medical Services, General Practitioners or Allied Health providers, such as dietitians.
   - Do you have any thoughts on what types of advice, resources, or referrals might be suitable, or not suitable?
   - If the health check were to provide advice or link you to referrals, how could this best be done:
     a) Advice or resources
     b) Referrals?
     Examples: SMS, Email, Posted, At the end of the health check online (printable/downloadable summary report).
   - Do you think a clinical review should be done first, before any advice or referral?
   - Do you think a phone call from a clinician is required to discuss results?

9. Regarding the questions I have asked you today over the phone: were they clear? Was anything confusing?

Note: For some multiple-choice questions, participant responses were coded to pre-existing response options based on what participants said, while for other questions, potential response options were communicated when the interviewer asked the question.
