## Appendix Three for "Preventive care in orthopaedic clinical services – testing the acceptability of an online health risk self-assessment tool using a multi-method design"

### Appendix Three- Qualitative feedback summary

The provided quotes are sometimes a combination of direct quotes from participants and the interviewer's interpretation of the feedback that was recorded during the consumer feedback interviews. This was required as notes were taken during interviews, which were completed via telephone and not recorded.

| Feedback Themes | Description (summary of theme) | Quotes |
|---|---|---|
| **Positive feedback/Enablers** | | |
| Design | User-friendliness-Tool was clear and easy to use | F, 50-59 years *'Thought it was quite clear, easy to navigate. Questions were clear and concise. Not information overload, good amount of questions and information'*<br>M, 30-39 years *'very easy to fill out. Impressed'*<br>F, 40-49 years *'easy to follow, was good, didn't have any issues.'*<br>F, 50-59 years *'Health check was clear, to the point, straightforward.'*<br>M, 60-69 years *'Was not too long. Short and sweet.'* |
| | Online convenience-SMS delivery and online tool was convenient and accessible | F, 50-59 years *'convenient to do online'*<br>F, 50-59 years *'Good to answer online, convenient'*<br>M, 20-29 years *'SMS simple and easy to access'*<br>M, 50-59 years *'SMS all doing it these days'* |
| | Motivational-Summary of answers based on national guidelines provided at the end of the health check was helpful and motivational | M, 50-59 years *'good for reminder of health factors'*<br>F, 50-59 years *'Planning to eat more veg'*<br>M, 50-59 years *'stop smoking motivation'*<br>M, 50-59 years *'Interesting to see the comparison, gives you an understanding of how you're going and where you are'*<br>F, 20-29 years *'realised need to have more fruit. Plans to try and change that'*<br>M, 60-69 years *'Motivational and informative... Was honest so got good feedback and got pat on back for things doing well.'*<br>F, 50-59 years *'Gave me a few different ideas, took it as positive feedback.'* |

| | | |
|---|---|---|
| | Helpful- Optional extra information on lifestyle risks, providing advice, resources and support service information was helpful | M, 20-29 years *'broad outline, good reminder of standards and importance to health'* |
| High importance | Participants understand their lifestyle factors are important to health and healthcare | F, 60-69 years '*very important, part of holistic approach to health*'<br>F, 50-59 years *'Good to eat healthy & give up smoking for surgery – faster healing'*<br>F, 40-49 years 'helps m*anage health in general'*<br>M, 50-59 years *'Important that health professionals understand the complete picture / can deliver holistic care, and share information with other clinicians where relevant'*<br>F, 60-69 years *'just retired after 40 years of nursing and thought the concept of preventive care was good'* |
| | Information was seen as important for improving health | M, 50-59 years *'Sometimes people don't realise they are doing something until it is in front of them.'*<br>M, 40-49 years *'Thought it was good information to have, hadn't seen that information before.'*<br>F, 50-59 years *'general information ok - knowledge is power.'* |
| Engagement with the health service | Appreciative of contact and care whilst on waitlist | M, 60-69 years *'Was good to get contact after long time on waitlist'*<br>M, 30-39 years *'Good to get questions'* |
| **Negatives feedback/Barriers** | | |
| Design | SMS delivery- Issues with appropriateness of SMS as delivery mode, including preference for alternative methods | F, 50-59 years *'Letter in the mail or a phone call would be preferred.'*<br>F, 60-69 years *'Might be better to send by email - can provide more information. Get lots of texts.'*<br>F, 60-69 years *'Not clever with that stuff* [online forms]*… mail would be better.'*<br>F, 60-69 years *'SMS may be difficult for older people - don't assume that people are computer literate'*<br>M, 30-39 years *'cultural understanding... understanding the cultural impact.'*<br>M, 60-69 years *'took time… not good with tech'* |
| | SMS wording was unclear, particularly | F, 60-69 years *'Not sure of the purpose.'*<br>F, 60-69 years '[needs] *more information on what it is about'* |

| | | |
|---|---|---|
| | purpose and instructions | F, 50-59 years *'I don't think it* [SMS wording] *made sense'* |
| | Belief that face-to-face preventive care by clinicians is more appropriate and personal | F, 60-69 years *'Prefer face-to-face to discuss in more detail'*<br>M, 60-69 years *'should be done face-to-face so that Dr can see patient, that way the truth can be seen*<br>F, 40-49 years *'face-to-face is the best time to discuss'*<br>M, 50-59 years *'prefer face to face for privacy and to get more specific detail'* |
| | Unable to complete-Technical difficulties or medical condition prevented completion | F, 50-59 years *'was just out of hospital… in recovery'*<br>F, 50-59 years *'recovery from shoulder surgery'* |
| | Question format/content-Uncertainty and uncomfortable with some questions that were asked, and questions/response options being limited | F, 20-29 years *'Confused as to why these questions were being asked'*<br>F, 60-69 years *'can't do exercise with bad knee, so found that question stupid... Offensive to ask about Indigenous status'*<br>F, 50-59 years *'Questions could have had more leeway... questions/responses were too cut and dried, could have had an opportunity to explain context (e.g. impact of condition)/responses a bit more'*<br>F, 50-59 years *'it's lifestyle and unavoidable, it can be upsetting.'* |
| | Need for inclusion of other questions-Interaction between mental and physical health, impact of pain and wait time on mental health, and suggestions to ask questions about mental health in health check | M, 60-69 years *'May be good to consult with mental health people about adding some questions that could help in this way'*<br>M, 30-39 years *'thinks this survey and the other online check could ask about mental health as this often overlooked and tied into the reasons for other risks such as smoking'*<br>F, 50-59 years *'add mental health and pain questions'*<br>M, 60-69 years *'Understand the reasoning* (for health check) *but I do not think it changes knee condition much... some psychology involved with pain. High degree of frustration because of condition and how disabling it is, has had suicidal thoughts.'* |

| | | |
|---|---|---|
| Risk | Concerns over online risk, including SMS links, security, privacy, legitimacy. Note: SMS alpha tag (unique sender ID) added to SMS in batch 3 (HNEHEALTH) significantly reduced concerns | F, 50-59 years *'Concerned initially about risk of opening -scams etc. Opened after second text'*<br>F, 60-69 years *'Good that it had logo (reduced concern about scams)'*<br>F, 50-59 years *'would be ok if know that the link is coming from a safe site'*<br>F, 50-59 years *'Wary of scams via SMS.'* |
| Engagement with the health service | Unexpected contact after years on waitlist either increased concern of SMS legitimacy or was an annoyance as there was no indication of an appointment | M, 60-69 years *'First contact in 2 years. Concerned about timeline'*<br>M, 60-69 years *'absolutely hopeless, contacted them twice to find out if was still on waitlist, SMS first correspondence'*<br>F, 50-59 years *'a phone call first would be best, so it is expected'*<br>F, 60-69 years '*Impacted life, not sleeping, can't work. I'm in pain, what I really need is a time-plan. Angry flying blind'*<br>F, 20-29 years *'may help to say in SMS not related to* (ortho) *appointment'* |
| | Unsure about potential impact of completing the tool on their orthopaedic appointment/care | F, 60-69 years *'I got the idea that responses would go back to ortho and potentially impact care/priority of appointment'*<br>M, 30-39 years *'Not likely to complete health check as it doesn't impact on wait list times.'*<br>F, 60-69 years *'Waste of time, as not linked appointment.'*<br>M, 50-59 years *'it doesn't get you closer to the surgery or the doctor'*<br>F, 50-59 years *'Got excited that survey related to ortho appt, disappointed to find out it wasn't.'* |
| Relevance | Not relevant to individual- Considered lifestyle health check to be not relevant due to being healthy, | M, 20-29 years *'Not relevant to self'*<br>F, 30-39 years *'unsure of benefit to self – healthy'*<br>F, 50-59 years *'good for people general info. not for self, doctor has helped and knows what needs to be done'*<br>F, 60-69 years *'Does not think needed as GP asks all these questions'*<br>M, 50-59 years *'survey doesn't tell you much and doesn't help'* |

| | | |
|---|---|---|
| | already leading healthy lifestyle, lifestyle risks already understood, or not interested | |
| | One size fits all approach- Concern that questions do not allow for personnel circumstance or extra detail, so that clinicians can give appropriate responses and advice | M, 50-59 years *'not specific to myself'*<br>F, 40-49 years *'most people receiving will have medical problems and don't want autobot response. Would prefer tailored response to medical condition'*<br>F, 60-69 years *'Summary - Not individualised enough'*<br>F, 60-69 years *'Responses to questions can be really influenced by condition/disability. Felt a bit awkward by the questions - felt that if someone was reading the answers that there might be some judgment, especially given very rare condition that is difficult for others to understand'*<br>F, 60-69 years *'Maybe start they survey with an opportunity to provide comment on existing conditions at the start, to provide context to responses'*<br>F, 60-69 years *'[needs] other languages options'* |
| | Low priority- Not completed because of other factors being more important | F, 50-59 years *'too busy'*<br>F, 20-29 years *'5 children and did not have a chance'*<br>F, 20-29 years *'didn't read the message'* |

Notes: SMS= Short Message Service (text message)
